# Hospice staff perspectives on referrals for specialist psychological support: a qualitative study across eight hospices in England

**DOI:** 10.64898/2026.11.25.26369007

**Authors:** Maria Valkovskaya, Richella Ryan, Elaine Mayon-White

## Abstract

**Background:** Psychological support in hospice care follows a stepped model in which all staff are expected to provide different levels of support to individuals under hospice care. However, the underlying reasons for referral to psychological specialists remain unclear. This study explored hospice staff’s perspectives on the referral process to psychology and developed recommendations to improve referral pathways.

**Methods:** This qualitative study used focus groups and individual interviews with hospice staff and applied an interpretative, reflexive thematic analysis approach. Sampling was purposive across eight hospices in England.

**Results:** Data from 52 health and social care professionals were included in the analysis: 48 from eight focus groups and four from individual interviews. Two overarching themes were developed. The first described the different ways healthcare professionals identified patients who needed more specialised support. The second focused on acting upon that need, i.e. making a referral to psychological specialists. The study highlighted a discrepancy between identifying patients with higher-level psychological needs and acting upon those needs through referral. Recommendations to improve psychological support provision include embedding in-house psychological specialists in all hospices, enabling quicker liaison with external providers, delivering training and supervision to staff involved in lower-level psychological interventions, and offering support and guidance for staff whose patients decline or are too unwell for referral.

**Conclusions:** Recognising a need for specialist psychological support did not always result in a referral. This study provides insights into hospice staff’s perspectives on referrals to psychological specialists and offers recommendations for improving referral pathways. Although the effectiveness of these recommendations was not assessed, psychological specialists and policy-makers can use the findings to inform improvements in psychological support provision in hospice care and evaluate the proposed changes in future research.

## Introduction

Hospice care is provided for people with incurable and life-limiting conditions [1]. It emphasises holistic needs, covering multiple domains, including medical, psychological, social, spiritual, and cognitive needs [2]. Unlike general palliative care, hospice care involves a specific hospice organisation directly delivering services, although the two terms are sometimes used interchangeably [3]. Hospice care prioritises quality of life and is associated with better patient experiences and outcomes compared to acute hospital admissions [4,5].

Psychological (or emotional) distress refers to a spectrum of emotional, cognitive, and behavioural responses to severe illness, loss, dying, and bereavement. In hospice and palliative care, some level of distress is anticipated and considered a natural response to serious illness and vulnerability [6]. This may include sadness, anxiety, fear, hopelessness, loss of meaning or purpose, and can interfere with an individual’s ability to cope [7]. Psychological support is a critical component of holistic hospice care [8], designed to address emotional distress.

The National Institute for Health and Care Excellence (NICE) guidance published in 2004 [9] described a four-level model of psychological support provision in UK hospices, with different levels of support depending on the complexity of patients’ psychological needs. This guidance was still in place at the time of data collection but has since been retired. According to this model, all hospice staff were expected to provide Level 1 psychological support, which includes basic emotional support, such as active listening and reassurance. Level 2 professionals are non-mental health specialists who have some additional training and can deliver low-level psychological interventions, such as stress management and relaxation techniques. Levels 3 and 4 include mental health professionals, such as psychotherapists, counsellors, psychologists and psychiatrists, who provide more specialised interventions for individuals experiencing higher levels of psychological distress.

Studies following publication of the 2004 NICE guidance [9] have described developments in psychological support provision in UK hospices, particularly at Levels 1 and 2 [10–13]. However, barriers remain, such as difficulties in consistently assessing and recognising psychological needs, and limited access to psychological specialists, i.e. staff working at Levels 3 and 4 [10,11]. These barriers were also identified in a recent mixed methods survey of hospice staff in the UK and Republic of Ireland, in which more than half of respondents reported unclear referral guidance and over one-third said they could not arrange appropriate psychological support when needed [14].

Considering the limited availability of specialist psychological support in palliative care, this scarce resource must be used effectively. Therefore, it is important to understand how hospice staff working at Levels 1 and 2 identify patients in need of more specialist psychological support, and what barriers they face when making specialist referrals.

Up until now, despite the prevalence of emotional distress in palliative care [15–16], research exploring the psychological domain of hospice care has been limited. A common limitation of existing studies in this field is that most have examined and classified referrals from the perspective of psychological specialists (e.g. [10,13,17]), while the voices of healthcare professionals making these referrals remain largely undocumented. The literature suggests that there might be some divergence in perspectives between psychological specialists and referring staff regarding referral reasons and urgency [10,13,18]. Qualitative research with hospice staff is needed to understand their perspectives on referrals to psychological specialists. Such insight can strengthen collaborative communication between psychological specialists and referring staff, and positively impact patients’ care [19].

## Methods

### Aim

The aims of this research were to explore the perspectives of hospice staff delivering lower-level psychological interventions on referrals to psychological specialists, and to develop context-specific recommendations for psychological support provision.

### Design

We conducted an interpretative qualitative study, underpinned by a critical realist ontology, which used a combination of focus groups (FG) and individual interviews with hospice staff. Data were analysed using reflexive thematic analysis [20]. The method was chosen as it enabled the exploration of patterned meaning in relation to referrals for specialist psychological support in hospice care. This approach also allowed space to recognise and engage with the researcher’s reflexivity as an integral and valuable component of the analytic process. We followed Braun and Clarke’s guidance on quality practice in reflexive thematic analysis [21] to support methodological rigour and the Consolidated Criteria for Reporting Qualitative Research (COREQ) [22] to guide reporting. The COREQ checklist is provided as S2 Checklist.

### Ethics and consent

Ethics approval was granted by the Psychology Research Ethics Committee at City, University of London (now City St George’s, University of London; reference: ETH2324-0154) on 7 November 2023. We also obtained local research approval from each site’s research governance committee or equivalent. Written and oral informed consent was obtained from all participants. All participants provided consent for publication of de-identified quotations.

### Reflexivity

This reflexivity statement is written from the perspective of the first author (MV), who collected and analysed the data, and was therefore most closely involved in shaping the interpretations presented:

I am a young woman who moved to the UK several years before starting my doctoral training. This study formed part of my Professional Doctorate in Counselling Psychology (DPsych) and therefore was conducted in my role as a trainee counselling psychologist. At the time of data collection, I already held BSc and MSc degrees in Psychology and Neuroscience from UK universities and had previous experience and training in conducting interviews and focus groups, gained during my two years working as a research assistant.

The reflexive component is central to this study. I consider myself to hold both an insider and outsider position in this research. My previous experience of working in a hospice setting positioned me as an insider, as I was familiar with the environment in which participants worked. One of the participating hospices was where I had previously completed a placement as a trainee counselling psychologist. Although my placement had ended by the time of data collection and I was no longer part of the psychology team, I knew some of the participants from my previous professional role. At the same time, I was an outsider because my background was in psychology, while the study excluded psychological specialists from participation.

I was drawn to hospice care through my own personal encounter with loss and the need to face my anxieties around death and dying. While on placement as a trainee counselling psychologist, I often encountered narratives about ‘inappropriate referrals’ to psychologists. Though anecdotal, these experiences inevitably shaped my analytical lens.

Encouraged by my research supervisor (EMW), I kept a reflexive journal throughout my doctoral research project to reflect on my assumptions, experiences and how these might influence my interpretation of the data. Following each focus group and interview, I recorded my reflections on the discussion, including my responses to participants’ accounts and any assumptions I became aware of. I revisited these reflections during the analysis to consider how my previous experiences and professional background might have influenced my interpretation of the data.

The pilot focus group was particularly important in helping me recognise how my professional background as a trainee counselling psychologist influenced my approach to facilitating discussions. Reflecting on the recording with my research supervisor (EMW) helped me become more aware of my tendency to adopt a therapeutic role when emotionally sensitive topics were raised and encouraged me to take a more facilitative approach in subsequent focus groups.

### Setting and sampling

The study was a multicentre project which aimed to recruit hospices from different geographical locations with different types of psychological support provision, and participants with diverse job roles. A purposive sampling strategy [23] was chosen. This sampling procedure did not aim for statistical generalisability but for analytical and theoretical generalisation [24]. The sample size was informed by previous research, which stated that three to six focus groups were needed to achieve saturation in focus group data, with four to twelve participants in each group [25]. However, this was used as a guide for initial planning only, and we did not aim to achieve data saturation, in line with the principles of reflexive thematic analysis [20]. All hospices that expressed interest in participating during the pre-planned data collection period were included in the study.

### Recruitment and data collection

All research-active adult hospices in England listed on the National Institute for Health and Care Research (NIHR) website [26] were contacted. All hospices which expressed interest in participating in the research during the pre-planned data collection period from November 2023 to April 2024 were included. Participants were recruited from nine hospices in nine geographical areas in England: Cambridgeshire, Coventry, West Midlands, Kent, Essex, South Yorkshire, East Sussex, Wiltshire, and West Yorkshire.

The researcher worked closely with research leads in participating hospices. Research leads distributed information about the study to hospice staff using an online leaflet and thus supported participant enrolment. The study introduced minimal exclusion criteria. Everyone whose job role involved the possibility of being involved in referrals for psychological support was eligible to participate. Staff whose main role involved providing specialist psychological support to hospice patients were excluded.

Focus groups with hospice staff were chosen as the primary data collection method to create a more ‘naturalistic’ environment for the participants, resembling a peer group discussion [27]. Individual interviews were reserved as an additional data collection method for consultant-level staff. This was to accommodate consultants’ time constraints and to mitigate the power imbalance between consultant-level staff and other employees. The researcher travelled to the participating organisations to conduct data collection face-to-face. Research leads were not present in the room during the data collection.

A semi-structured topic guide was used for both focus groups and individual interviews. Questions were adapted depending on the flow of the discussion. Before the main data collection, a pilot focus group was conducted in one of the participating hospices to assess the clarity of the questions and whether they addressed the research aims. The pilot followed the same recruitment and consent procedures as the main study. Following the pilot, the audio recording was reviewed and discussed in research supervision. This highlighted that the researcher had sometimes adopted a more therapeutic rather than facilitative role, particularly when emotionally sensitive topics were discussed. In addition, the researcher tended to direct the conversation to ensure that all questions were covered, which resulted in the focus group lasting longer than planned. Following this reflection, the researcher adopted a more facilitative approach in subsequent focus groups, encouraging participants to respond to one another and allowing the discussion to develop more naturally. As the facilitation style of the pilot differed from that of the subsequent focus groups, the pilot data were excluded from the final analysis.

The topic guide, participant information, consent forms and researcher’s introduction are provided in S1 Appendix (sections A-D). The written introduction described the researcher as a counselling psychology doctoral student and explained the study purpose. All focus groups and interviews were audio recorded with participants’ consent using a secure recording device. No field notes were taken during data collection, but the researcher recorded reflections in a reflexive journal immediately after each focus group or interview. Transcripts were not returned to participants for correction or comment. Focus groups lasted between 58 and 76 minutes, while individual interviews lasted between 28 and 41 minutes.

### Analysis

All focus groups and interviews were audio recorded and transcribed verbatim. Transcripts were coded manually using the comments functionality in Microsoft Word. Data were analysed using reflexive thematic analysis by Braun and Clarke [20]. MV coded transcripts and generated the initial codes and themes. These were discussed with EMW throughout the analysis to explore different interpretations, reflect on assumptions and support the methodological coherence of the analysis. A predominantly inductive orientation was chosen being particularly suitable for under-researched questions. Following Byrne [28], both semantic and latent coding were used. Latent coding involved looking beyond the surface meaning of the data and considering possible assumptions or ideas underlying what participants were saying. This helped to deepen the semantic coding by adding another layer of interpretation.

Reflexive thematic analysis involved six phases: familiarisation with data; coding; generating initial themes; developing and reviewing themes; refining, defining and naming themes; and writing up [20]. These were not linear but involved forward and backward processes and awareness of personal and epistemological reflexivity in all the stages. All phases were overseen by EMW.

### Patient and public involvement

Patients and members of the public were not involved in the design, conduct, reporting or dissemination plans of this study.

## Results

### Participants

Nine focus groups (including one pilot group) and four individual interviews were conducted between January and April 2024. The data from the pilot group were excluded from the analysis. Therefore, all results presented below are based on the main study involving eight focus groups and four individual interviews across eight hospices in England. The number of participants in each group varied from three to eight individuals, depending on the number of people who expressed interest and was also likely affected by the size of the hospice and their ability to release staff from duties. As focus groups were the primary data collection method, at least one was conducted in each participating hospice. Each of the four individual interviews was conducted in a different hospice.

Data from 52 participants were included in the analysis, with 48 participating in focus groups and four in individual interviews. Table 1 provides an overview of participant characteristics. Demographic characteristics and job roles were self-reported by participants. The majority of participants were female (n = 45). Participants’ experience in hospice care ranged from 1 month to 34 years, with most having worked in hospice settings for over 5 years. Participants included a range of professionals, such as doctors, nurses, social workers, physiotherapists and support staff. To reduce the risk of identifying individual participants, some job roles were grouped under the ‘Other’ category in Table 1. There were no missing data for the characteristics presented in Table 1.

**Table 1.** Participant characteristics (n = 52).

| Characteristic | Category | n (%) |
|---|---|---|
| Job title | Doctors | 9 (17) |
|  | Nurses | 20 (38) |
|  | Social workers | 8 (15) |
|  | Physiotherapists | 2 (4) |
|  | Occupational therapists | 3 (6) |
|  | Complementary therapists | 3 (6) |
|  | Support workers | 3 (6) |
|  | Chaplains | 1 (2) |
|  | Others | 3 (6) |
| Years working in hospice care | ≥ 5 years | 35 (67) |
|  | < 5 years | 17 (33) |
| Participants’ sex | Male | 7 (13) |
|  | Female | 45 (87) |
| Participants’ ethnicity | White | 50 (96) |
|  | Other ethnic groups | 2 (4) |

Percentages are rounded. The experience summary reports the proportion with more than five years in hospice care.

Table 2 outlines the structure of psychological support provision in the participating hospices (n = 8). Three hospices had in-house access to Level 4 staff, five to Level 3 staff, and two reported no in-house access. Because some hospices had access to both Levels 3 and 4, the total shown across categories exceeds the overall sample of eight.

**Table 2.** Psychological support provision at participating hospices (n = 8).

| **Psychological support provision** | **n (%)** |
|---|---|
| In-house access to Level 4 staff | 3 (38) |
| In-house access to Level 3 staff | 5 (63) |
| No in-house access to Level 3 or 4 staff | 2 (25) |

Categories overlap because some hospices had access to both Level 3 and Level 4 staff. Percentages are rounded.

### Thematic analysis results

Two overarching themes and seven subthemes were developed (Table 3). The first overarching theme describes the process of identifying the need for specialist psychological support, while the second explores acting upon that need, i.e. making referrals to psychological specialists.

**Table 3.**
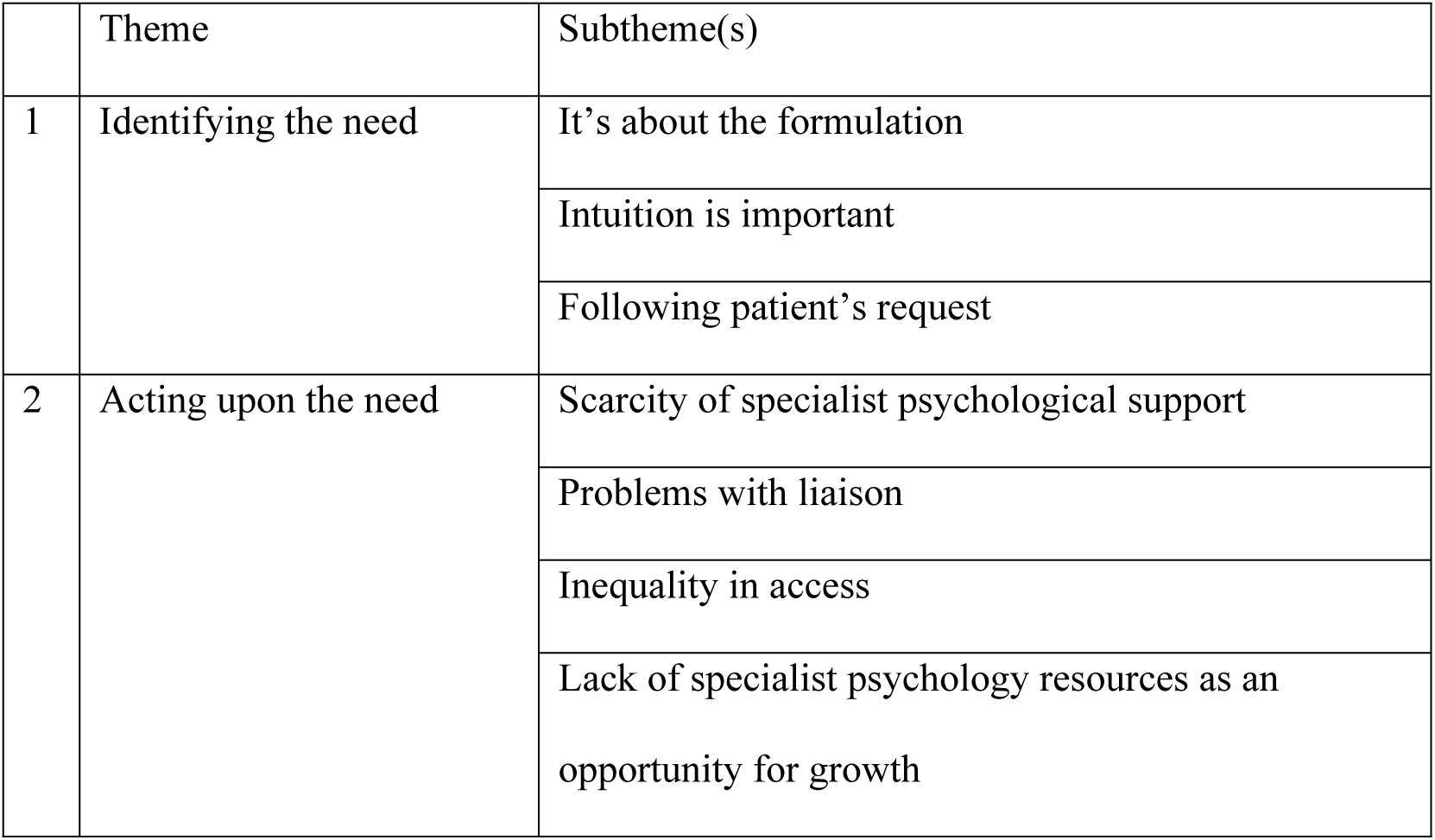
Themes and subthemes.

|  | Theme | Subtheme(s) |
|---|---|---|
| 1 | Identifying the need | It’s about the formulation |
|  |  | Intuition is important |
|  |  | Following patient’s request |
| 2 | Acting upon the need | Scarcity of specialist psychological support |
|  |  | Problems with liaison |
|  |  | Inequality in access |
|  |  | Lack of specialist psychology resources as an opportunity for growth |

Participants’ quotations are presented using pseudonyms.

#### Theme 1. Identifying the need for specialist psychological support

In this theme, we explore how participants described their thinking process around referring patients for specialist psychological support — from an analytic approach involving symptom assessment, questionnaires, and team discussions, to a more intuitive approach based on gut feelings, and, in some cases, responding directly to patients’ own requests for referral.

### It’s about the formulation

Formulation is a core skill of practitioner psychologists in the UK [29–31]. It refers to how psychologists make sense of the causes of people’s distress and use this understanding to inform further intervention [32]. Formulation may occasionally be employed by other health and social care professionals, particularly within mental health contexts, although the term itself is not always explicitly used [33–35]. I named the subtheme It’s about the formulation to reflect my interpretation, as a trainee counselling psychologist, of how hospice staff engage in a process of making sense of patients’ psychological and emotional presentations. The name of the theme, therefore, reflects the duality of the data in line with the Reflexive Thematic Analysis which acknowledges the researcher’s subjectivity as a useful resource that needs to be considered rather than bracketed [20].

Participants demonstrated a strong awareness of the four-level model described in the NICE (2004) guidance [9], which was still in place at the time of data collection but has since been retired. However, they reported a lack of more specific guidance on when patients should be referred to psychological specialists.

Hospice staff emphasised the need to consider many different factors when deciding upon the level of psychological needs. Participants described the importance of considering the patient’s situation holistically, i.e., examining multiple factors, including the patient’s medical needs, social needs, prognosis, support system, and their resilience level. As such, it appeared that the thought process was fluid rather than something happening at a particular point. As Tracy (FG6) put it:

> And I think as well as a multidisciplinary team right through the process from the first assessment when people first come onto our service through the, the environment in the community, when they’re having contact with the nurses and other professionals, and then if they come on to the inpatient unit, we’re all constantly assessing. And as part of that, we’re kind of building up a picture and hopefully a holistic picture of, you know, who they are, what their strengths are, what their support network looks like, where the vulnerabilities might be, er and aiming to kind of address those as we can. And sometimes if we’re finding a gap, we try to sort of fill it. So hopefully by the time people get to a point where things are going wrong, we have a sort of an understanding of who they are. It’s not always the case, but it gives us a baseline to kind of make decisions from.

As illustrated in the quote above, participants repeatedly emphasised the use of the multidisciplinary team (MDT) in the formulation process by ‘bouncing ideas’ and considering ‘a whole multitude of factors’ (Harvey, Interview 1). Communication between different team members was consistently highlighted as important, particularly in relation to the continuous and evolving nature of formulation, that is, the analytic process involved in evaluating the patient’s needs.

When speaking of identifying the patients with higher level of psychological needs, participants often talked about ‘complexity’. For many participants ‘complexity’ meant ‘something else’ on top of the palliative diagnosis, as Sara (FG5) illustrated in her formulation process:

> I suppose it’s that, is this a palliative care need as well as an X?

To define the complexity, participants frequently spoke about ‘red flags’ which I understood as factors that would increase the probability of a higher level of psychological needs. In Table 4 I have summarised the patterns in the ‘red flags’ that participants often mentioned across all focus groups and interviews. However, in line with reflexive thematic analysis, I deliberately avoid stating frequencies to prevent the impression that frequency determines importance [20,36].

**Table 4.** Referral considerations described by hospice staff.

| **Category** | **Reported reasons for referrals** |
|---|---|
| Risk and Crisis Management | Suicidal ideation/self-harm, suicide attempts, challenging behaviour, domestic abuse, safeguarding needs, staff risk. |
| Mental Health Conditions and Psychiatric Needs | Pre-existing psychiatric diagnoses, symptoms suggestive of post-traumatic stress disorder, agoraphobia, cognitive impairment, substance abuse, mental health symptoms triggered by palliative diagnosis without previous psychiatric history |
| Psychological Distress Impacting Physical Health | Psychological distress as a barrier to treatment, exacerbating physical symptoms, anxiety worsening health, psychological manifestations of distress (pain, agitation), 'total pain' concept, CBT for chronic conditions |
| Preventative and Supportive Referrals | Anticipating difficult transitions, supporting parents to help children cope, learning coping strategies, escalating carer distress. |
| Severe or Prolonged Emotional Distress | Withdrawal from care team, excessive crying, rumination, complicated grief, inability to cope, ‘inappropriate’ reactions, distress-prone groups (young patients, sudden diagnosis, motor neuron disease patients) |
| Psychosocial and Relational Challenges | Complex family dynamics, adjusting to changes in relationships, multiple losses, high external stressors |

Referrals reasons were coded and interpreted in context. CBT stands for cognitive behavioural therapy.

One of the ‘red flags’ that was brought up in all focus groups and individual interviews was patients talking about suicide or a wish for assisted dying. This was clearly an uneasy area for the participants to discuss, as was manifested by their verbal reactions, long pauses and sighs (FG4).

> Katie: Like a suicide?
>
> Lisa: Yeah, oh God, yeah
>
> Sarah: ((sigh))
>
> (…)

Participants felt that mental health history (either self-reported by patients or documented in the care records) would be another common red flag contributing to complexity. For example, Kushal (FG7) explained:

> That is actually, that actually automatically rings alarm if someone is already depressed or anxious, so then they are in such a difficult situation. How is it affecting them?

Nevertheless, the participants argued that none of the ‘red flags’ taken in isolation would automatically imply complexity; rather, it was the process of formulating a holistic picture of the person.

They also felt that, on many occasions, screening tools they used (e.g., questionnaires) supported the formulation they had already constructed. However, participants were clear that the score alone was never enough to justify referral for higher-level psychological needs; instead, it served as a prompt for deeper exploration of the issue.

> So erm, if somebody scored very highly so as you know, showing high signs of distress, we, it would, I don’t think it would be the only factor. It would prompt us talking to the patient a bit more about it, and that would then feed in. (Harvey, Interview 1).

### Intuition is important

Alongside the formulation process, which involved a careful weighing of different variables, participants emphasised the importance of trusting themselves and used words such as gut instinct, intuition, and sixth sense to describe this feeling. Many strongly believed that ‘gut instinct’ was the most important criterion in identifying whether a patient or their close relatives might have specialist psychological needs.

Interestingly, participants described how this feeling could emerge even before any conversation took place, as if they could sense a shift in the atmosphere in the room, yet struggled to pinpoint exactly what it was.

> There are some rooms when you go in where it is palpable. (Lisa, FG4)

Healthcare professionals working in hospice care were very aware that many, if not most, of their patients would present with a level of distress. However, what was described as ‘gut feeling’ was crucial in helping them find the fine line between what they described as ‘normal’ and ‘abnormal’ distress. It seemed as if intuition played a vital role in enabling hospice staff to hear beyond words and understand what their patient was expressing. This required fine attunement, mastery of listening skills, and curiosity, that is, a willingness to see beyond what the patient was saying with their words. It felt as though important information could have been easily missed if staff relied solely on analytical thinking, without paying attention to their inner sense that something was ‘not quite right’ with their patient. Phoebe (FG3) explained:

> Phoebe: And I think on the telephone, I think this is where sometimes it goes wrong because people think, ‘Well, they’re not really saying a lot erm let’s move on’. And yet the patient is crying out or a relative is crying out and then they think you haven’t listened. But actually, they
>
> Elizabeth: Really hard

Hospice staff emphasised the role of learning from experience, which played a crucial role in helping them develop gut feeling and the ability to trust it. Many reflected that they would have struggled to put into words the ‘objective criteria’ for describing the ‘abnormal’ but could easily sense it and make quick judgements based on their intuition. Leo (FG2) commented:

> And then that just becomes sort of (.) the (.) your your standard deviation of what is it no-what is a normal response to terminal illness and people who drift outside that. That’s a very hard concept to explain to someone who’s not had lots of experience before (..).

Even though participants referred to the process of relying on feeling to identify specialist needs as ‘subjective’, this did not mean unreliable. Hospice staff described how their colleagues would often agree on the same feeling regarding a patient, sensing that something was not quite right.

> I think it’s easier on the ward because obviously you’re. They are there in front of you. And we bounce off each other, don’t we, so, you know, you go to the room, come out and then you say to your colleagues ‘Oh, do you know what? I’m really, really worried about, you know, patient number four’. And they’d also ‘Actually, I felt that, too’. (Mia, FG3)

It seemed that instead of choosing between the two (analytic OR intuitive), participants were suggesting that while, on a day-to-day basis, they valued and trusted their intuition, it would also be helpful to have some criteria to lean on, especially to facilitate the learning of newly qualified clinicians.

> I suppose having some kind of guidelines that other people have experienced as well, that’s written down. So when you do have somebody new, it’s kind of, it’s not, it’s like, you not just say, ‘It’s just a feeling that something’s not quite right’. If we can actually voice it or write it, it would be a good way for somebody who’s new to say, ‘Oh, yes’ (Maya, FG8)

Thus, while valuing intuition highly in identifying specialist psychological needs in the patients, participants were also able to reconcile it with analytical thinking and see how both could complement each other. Clinicians recognised that intuition and the analytic thinking could coincide, inform, and support one another, working together to provide the best possible care for the patients under the hospice care.

### Following patients’ requests

A patient’s unprompted request for psychological support was the third factor playing a role in identifying the need for a referral. Many participants noted that it was not uncommon for patients to identify psychological needs in themselves and therefore ‘request counselling.’ The request would often be passed directly to a psychological specialist, representing a ‘short-cut’ compared to a need identified by a healthcare professional, which would usually involve a formulation process and team discussion. Here, Daisy (FG5) describes how she would follow the patient’s request:

> If they specifically say they want some counselling, then that would, I would, I would kind of I would then go and talk and say, ‘You know, they’ve specifically asked for counselling, not just support or some or, or appeared to be in need, but they were specifically looking for something further’. And then I’d go and talk to them [psychological specialists].

The participants were certain that they would take the ‘self-identified’ need for specialist psychological support seriously and would follow up on the request to ensure it was addressed. Many also emphasised the value they placed on the self-referral process as this indicated that the patient was active in their own care and therefore more likely to engage meaningfully with the therapeutic process.

> And also people can be encouraged to self-refer because I mean, we have leaflets and part of what we’re sort of moving towards is giving people the information and then letting them make an informed choice about whether to request that service at that time or actually to come back to it at some other point if they need it. Because in doing so, they’re kind of cementing the commitment to the process. So if they actually self-refer, they are going through, they are walking the walk as well as talking the talk. So that’s one element of it. (Amy, FG6)

This excerpt also demonstrates how the patient’s agency in hospice care is promoted and how the patient is encouraged to take charge emphasising the philosophy of hospice care as person-centred.

#### Theme 2. Acting upon the need: ‘There is a need, but now what?’

The current theme explored the role of the system in the referral process. Rather than acting in a vacuum, patients and healthcare professionals were embedded within a broader system, which inevitably influenced their interactions. This theme suggests that some referrals to psychology were omitted not because the patient did not require psychological intervention but due to the absence of psychological specialists (i.e., resource limitations within the system).

### Scarcity of specialist psychological support

The desire for more specialist psychological resources for patients was raised in all focus groups and interviews. Participants often spoke about the demand for specialist support exceeding capacity and their wish to be able to refer more patients:

> So if I have, if I had a magic wand, I would want more clinical psychologists in palliative care, definitely, because I see clinical psychology is quite a precious resource and we have to allocate to those that really, really need it. But I probably see other people where they may benefit. […] So, really, if I could make one change, it would be more. I think every hospice should have a clinical psychologist who’s a member of the MDT. Both to support the patients, but also to encourage and empower the staff in providing that support. (Harvey, Interview 1)

Due to the scarcity of specialist psychological support, hospice staff felt a strong need to advocate for psychological needs to be taken seriously and for patients to receive support appropriate to their level of need. Paula (FG1) explained:

> I’m just thinking about like definition of palliative care, you know er I can’t remember exactly what definition is. It’s a few, but it talks about the we and again, the hospice mission that we’re addressing people’s physical and psychological well-being. So, so I know, like we say, like you feel like you have to sell a story er for someone, to er but it’s actually it is, it is equally important. I think it depends on probably erm leadership management at other levels to remember that physical and psychological are both the same. You can’t have one without the other.

Many complained about the pressures imposed on them by stakeholders and the system in its current form. Participants felt that the system was more preoccupied with adherence to formal requirements rather than being holistic and person-centred. Quite a few expressed their dissatisfaction with such a striking shift away from the values of the hospice movement in its initial form. Healthcare professionals felt that these processes impeded MDT discussions, making them less holistic and disrupting the formulation process. Participants in FG5 shared:

> Dave: I think sometimes the MDTs are taken over by erm
>
> Daisy: the time limits
>
> Dave: like process of it
>
> Daisy: process and the time limit
>
> Dave: I haven’t been to the inpatient unit and weekly MDT for ages, but that’s kind of like, ‘Have you ticked these various boxes? Has the IPOS been done? What’s the’
>
> Daisy: basically
>
> Dave: as opposed to a bunch of professionals get together and say, ‘Oh, you saw them, but were you talking about mental health stuff? What was that all about? Oh, right. Okay. That’s your’. Yeah, that’s sort of informal discussion as opposed to what’s
>
> Daisy: ‘Have you ticked this box?’
>
> Anna: That would actually Identify the gaps better, I think, than a tick box exercise

Participants, therefore, felt that difficulties with making referrals were often related to limited resources and the way services were organised.

### Problems with liaison

With regard to external services providing specialist psychological support, staff expressed frustration about the mismatch between the sense of urgency within hospice care and the longer waiting times typical of services outside the hospice. Hospice staff tried to articulate this to external services but they were often left feeling unheard. As William (Interview 3) put it:

> Yeah, yeah, yeah. And of course you know the IAPT system or I think now it’s called Talking Therapies. It’s too, it’s too, it’s the time frames are too prohibitive and I don’t think that kind of geared up for the things that we’re referring to. So that, that, that is not, erm, you know, that’s not really appropriate for our group.

As a consequence of being unable to access psychological specialists externally, hospice staff described how they had to actively problem-solve to meet patients’ needs using their own internal resources. This, however, had a significant and challenging impact on many members of staff involved in patients’ care. This was summed up by FG3 participants, who commented:

> Elizabeth: So we can say it’s urgent but they don’t. Because they are external agency, they don’t always tell us what their waiting lists are like. So erm, the communication is just not as good as it could be. So it’s really tricky because we say it’s urgent. But yeah, like I say, we then don’t, yeah. What time frame they’ve got. So I think it’s then often ends up with someone else in the well-being team picking it up.[…] It was difficult
>
> Mia: Very, very difficult. Difficult for the staff as well (.)

### Inequality in access

Notably, quite often there was a sense of inequality within the system, as access to specialist psychological support depended on geographical location, the patient’s diagnosis, and the type of care provided (inpatient or outpatient).

> Ella: And probably support for all, not just for the cancer patients really.
>
> All: ((General agreement))
>
> (FG7)

Out of all the patients, participants expressed concern that the system was often discriminatory towards hospice patients with complex psychiatric needs. Participants felt that hospice patients with psychiatric needs were effectively written off the records, with the door shut on any opportunity for shared holding between the psychiatric and palliative care systems. Beyond the impact on patients’ quality of life, reported difficulties with obtaining psychiatric referrals meant that hospice staff were left to find alternative ways to address patients’ unmet specialist needs, even when the recommended course of action was referral to a higher tier of care. This meaning was captured by FG8 participants:

> Jenny: So with the community psychiatric team, that’s quite tricky to actually get and to accept patients that they seem to be really reluctant to take patients who have got a terminal diagnosis because they their perceptions is there’s not it’s not worth, I don’t know, I can’t answer for them, but they seem to think that it’s that it’s not so valid to follow up a patient who’s got a very limited life expectancy. And that again, I’m just surmising sometimes I get the feeling that they don’t feel confident or don’t know what to do about patients who are approaching the end of life. So they rather rather then just say, ‘No, it’s not relevant for us to get involved’ yeah
>
> Maya: That was quite disheartening […]
>
> Jenny: And it is, it is frustrating because it’s, that’s never the first resort to ask them to get involved. It’s when we as a multidisciplinary team exhausted all the ideas we can come up with usually, or unless it’s blatantly obvious that it’s a psychiatric. Particularly with people with a longstanding history. But to then sort of hit a brick wall and, and, you know, sometimes they’ve literally put their foot down and say, you know, ‘That’s not appropriate for us’, and then what we meant to do?

Talking about the psychiatric patients, practitioners usually referred to the most complex patients with years of psychiatric history prior to their terminal diagnosis. Consultant-level staff were clear that they needed liaison with external psychiatry services, not for the prescription of the psychiatric medications, which palliative care consultants usually felt confident at, but it was the continuous support (the therapeutic part) that was missing:

> And he’s got quite a lot of anxiety and panic, you know, so a lot of it is cognitive worry about future and you know what’s going to happen to him and decision-making and but there’s also just sheer panic about going out like going out in outside of the house. Now that would have been somebody we would have referred to the mental health practitioner for a kind of acceptance and commitment cognitive approach, behavioural approach […] A lot of the psychiatry, though, I have to say, we can do. You know, what I mean? Like if it’s making a diagnosis, if it’s making, if it’s them prescribing antidepressants, it’s relatively straightforward. It’s, you know, prescribing benzodiazepines, it’s relatively straightforward. If it’s prescribing antipsychotics, we’ve got a good handle on that. So that meant that the kind of medical bit feels relatively straightforward. (William, Interview 3)

It was as if psychiatric patients in hospice care ended up stuck in a system that strongly favoured dichotomies and clear categorisation, leaving no room for intersectionality, such as being both a palliative patient and a psychiatric patient. This drive for simplicity and reluctance to communicate and provide shared care meant that the most complex patients were often left between services, without appropriate ways to meet their needs.

Interestingly, the confusion often arose at the entry point when people with palliative diagnoses were considered complex due to their psychiatric needs and were referred to local hospices that were unable to meet their needs due to the aforementioned problems with liaison and communication. This was summed by Jenny (FG8):

> I think what we find, as well as GPs aren’t clear about who’s responsible for what aspects of the psychological or psychiatric support, and now often refer to us when it’s much more of an obviously primarily psychiatric situation.

These hospice care referrals, therefore, created significant back-and-forth communication between different settings from the very start, using up valuable resources without making any real impact on improving the person’s quality of life.

Overall, the lack of an established pathway for referring hospice patients to external services, along with differences in hospice structure, contributed to inequality in the provision of specialist psychological support for hospice patients. It also highlighted a mismatch between the number of patients with specialist needs and the available resources to meet those needs, thus identifying key areas for improvement.

### Lack of specialist psychology resources as an opportunity for growth

At the same time, a lack of specialist psychological support readily available for all categories of hospice patients also created an opportunity for growth for hospice care professionals. For example, there was a strong pride in getting upskilled and liaising more with other internal teams to meet the needs of complex patients. This experience of professional growth was especially strong among staff working in hospices without any in-house specialist psychological support available. Some participants believed that they had learned to address specialist needs with resources available within the team and that patient care was not compromised. For example, Lisa (FG4) expressed:

> And I, don’t you think, that because actually we don’t have that psychologist in the day hospice because to me there’s not that resource there anyway. Therefore, we’re using the multidisciplinary holistic approach, which we’re doing. And we might not know the scientific reasons why it’s working, but we’re all kind of chipping away. And you know, I, I’ve been here [number] years, and I haven’t known anyone who, a handful of people that have died psych, you know, in a lot of psychological distress. I mean, yeah.

The question which arose was as to whether psychological specialists were an essential part of hospice care provision and if meeting the needs of patients with the already available resources within the MDT was a more cost-effective process. Although at first glance this appeared to be an attractive option, the same healthcare professionals were vocal about its downsides, particularly the impact on staff. As mentioned before, in-house psychological specialists often played a vital role in supporting staff as well as delivering direct interventions to the most complex patients. Therefore, while hospice staff working at Levels 1 and 2 did their absolute best to support patients, they had to constantly stretch themselves, without being given adequate time or support to recover. As Pauline (FG4) put it:

> I suppose I’m fortunate I have a psych, I have clinical supervision once a month, so I actually have somebody who does have that skill to help me, but unfortunately somebody who’s band five and going in and out of that and providing the level of care that you provide someone, you don’t have access to that level of clinical supervision that I have access to. And I think that’s a real shame and a real need.

## Discussion

### Key findings

This study presents a multidisciplinary account of referrals to psychological specialists from the perspective of hospice staff delivering lower-level psychological interventions.

The study found a discrepancy between identifying patients with specialist needs and acting upon those needs, that is, making a specialist referral. The latter was heavily dependent on the available resources within the system. Referral behaviours appeared to reflect interdependent structural and cultural dynamics rather than isolated clinical decisions.

The study demonstrated that the reasons underlying referrals to specialist psychological support extend beyond a simple list of patients’ diagnoses, presentations, or behaviours. The findings of this research suggest that the reasons underlying referrals are complex and involve the interplay between the patient’s symptoms, the skillset of healthcare professionals around the patient, and systemic factors, such as the availability of specialist services.

Overall, there did not appear to be any definite cut-off criteria indicating when a referral to psychological specialists should be made in any of the hospices that participated in our research.

The system was fluid and shaped by systemic factors, such as the availability of resources, as well as patient-related factors.

### Implications for policy and practice

Several findings were made which might be applicable for clinical practice and policy.

Firstly, our study demonstrated that reasons underlying referrals are more complex and nuanced than those presented in previous research where referrals were based on pre-assigned categories defined by psychologists, rather than referring clinicians (e.g. [17,18]). Although it might be necessary for audit purposes to classify referrals into fewer categories, it is also important for psychological specialists, managers, and guideline commissioners to recognise the complexity of presentations and the variability of reasons behind the neat categories. These can also provide insight into the training needs of staff working at Levels 1 and 2 by highlighting the presentations hospice staff consider in need for referral to a higher tier.

Secondly, in our study participants expressed that they tried to accommodate patients’ requests to see a psychological specialist by making specialist referrals. This aligns with current NICE guidance, which emphasises involving patients in decisions about their care and taking their wishes and preferences into account [37]. This also highlights that some referrals to psychological specialists might reflect patients’ wishes rather than the complexity of their presentations as identified by hospice staff working at Levels 1 and 2.

Thirdly, our study supports previous findings of a persistent gap between the need for referrals and the availability of psychological specialists. In the absence or shortage of in-house psychological specialists, the 2004 NICE guidance [9] described liaison with external services. Our participants often expressed uncertainty about referral pathways and the availability of external resources for their patients, especially the most complex and vulnerable adults, such as with existing psychiatric diagnoses and safeguarding issues. Our findings also align with the quantitative findings reported by Zavery et al., which highlighted unclear referral guidance, limited access to mental health specialists, and difficulty arranging appropriate psychological support when needed [14]. These accounts can also be considered alongside Stevens’s review of leadership and safeguarding vulnerable adults [38], although the present study cannot establish change over time. In the 2021 survey, approximately half of responding UK hospices reported relying on external referrals to access Level 4 staff [10], i.e. psychologists and psychiatrists – those whose expertise is required in the most complex cases. Our findings, therefore, echoed the challenges identified by Paley et al. [11], Russell and Fountain [13], and McInnerney et al. [10], including long waiting lists, lack of in-house specialists, difficulty accessing external referrals, and insufficient funding for specific groups of patients such as those with non-cancer diagnoses.

Stevens [38] suggests that change needs to be implemented on three levels: the societal level (broader socio-political and economic factors affecting vulnerability), the organisational level (staffing, policy, and governance), and the personal or individual clinical level. The four-level model described in the 2004 NICE guidance [9] provides historical context for this study.

Although the NICE (2004) guidance has since been retired, our findings remain relevant to psychological support provision in hospices. They highlight that identifying patients with specialist psychological needs is only one part of the referral process, as the availability of resources also influences whether these needs can be addressed.

The findings of this study suggest that change needs to be initiated at the policy-maker level to consistently embed the work of psychological specialists in local hospices, or, at the very least, to establish quicker referral pathways for hospice patients to existing external services. While drawing on these findings to provide recommendations for improving psychological support provision in UK hospices, I believe that offering isolated training to staff working at Levels 1 and 2, aimed at enhancing their assessment skills, without ensuring a sufficient workforce at Levels 3 and 4 (i.e., at the specialist level), is unsustainable and places additional pressure on staff who are already overstretched.

A summary of recommendations based on the researchers’ interpretation of the data collected during the study, as reflected in the themes and codes developed during the analysis, is provided below. A more detailed set of recommendations is included in S1 Appendix, section G. These recommendations reflect the researchers’ views on possible ways to improve practice and would need to be adapted to individual services. Their feasibility, effectiveness and cost-effectiveness in clinical practice have not yet been evaluated.

#### Recommendations for Service Leads and Policy Makers

- Advocate for increased funding for specialist psychological support provision by acknowledging psychological specialists’ dual role in supporting complex patients and staff working at Levels 1 and 2 by providing supervision and training.
- Advocate for the provision of specialised psychological support for all patient categories.
- Explore timely access to psychiatric liaison, including locally appropriate on-call arrangements.
- Provide an in-house psychological specialist to address gaps in psychological intervention and support for Levels 1 and 2 staff.
- Advocate for record sharing to ensure continuity of care.

#### Recommendations for Psychological Specialists

- Provide feedback to the referrer about their referral.
- Give updates regarding your team’s waiting list.
- Offer opportunities for further training in psychological skills.
- Support staff who assist patients who have declined a referral or are not well enough for one.
- Consider asking the patient for consent to share aspects of your work with the wider team, which will facilitate psychological support beyond your sessions.

#### Recommendations for Staff Working at Levels 1 and 2

- Acknowledge the work you are already doing.
- Explore the psychological skills and perspectives of your colleagues.
- Consider involving other teams, apart from psychological specialists, to address psychological distress.
- Consider making informal inquiries before submitting a referral.
- Seek support from psychological specialists to enhance your skills.

### Limitations and strengths

This study utilised qualitative data collected from a multidisciplinary sample of staff with varying levels of experience across eight hospices. To the best of our knowledge, this is the first study that explored referrals to psychology from the perspective of hospice staff making the referrals. The research highlights the complex reasons underlying referrals to psychological specialists, and their interplay with systemic factors influencing referrals. Based on these findings, a set of recommendations has been developed to enhance clinical practice and promote a more integrated approach between psychological specialists and medical staff.

The study has several limitations. Although participants were recruited from eight hospices across England, only research-active hospices that expressed interest in the study were included. Therefore, the findings might not reflect the experiences of staff working in other UK hospices. Recruitment through research leads and staff availability might also have influenced who took part in the study. As focus groups were conducted with staff working in the same hospice, existing relationships and differences in professional roles might have influenced the discussions. In addition, individual interviews were only conducted with consultant-level staff, making it difficult to know whether differences in participants’ accounts were related to their roles or the method of data collection.

The study explored hospice staff’s perspectives on referrals to specialist psychological support rather than observing actual referral decisions or measuring patient outcomes. Only two participants were from ethnic groups other than White, and limited demographic information was collected. Therefore, it is unclear whether the participants reflect the wider hospice workforce. Patients and family members did not take part in the study. Future research should explore their experiences of how psychological needs are identified, discussed and addressed, as well as their experiences of accessing specialist psychological support. Finally, although recommendations were developed based on the findings, they reflect the researcher’s interpretation of participants’ accounts, and their feasibility, effectiveness and cost-effectiveness have not yet been evaluated.

## Conclusions

Our study provided insights into referrals to specialist psychological support, an under-researched aspect of hospice care in England. It showed that hospice staff identified patients for referral to a higher tier through a combination of interpreting the patient’s symptoms (analytic process), relying on intuition informed by their understanding of what is considered ‘normal’ in hospice care, and listening to patients’ wishes to be referred. The findings also revealed that actual referrals were strongly influenced by the availability of psychological specialists within the system.

This research illustrates the importance of looking beyond the individual healthcare professional or patient when considering the reasons behind referrals to psychology. In some cases, where resources were available, healthcare professionals referred patients simply because they could; in other instances, referrals were not an option despite clear need. Although hospice staff demonstrated determination to provide the best care for their patients within very limited resources, it is essential to recognise the impact of the wider system on hospice care, beyond individual factors.

## Data Availability

The full transcripts of the focus groups and individual interviews are not publicly available due to the potentially identifiable nature of the data. Although identifying information was removed, some details could make it possible to identify individual participants or hospices. De-identified quotations supporting the findings are included in the manuscript. Additional de-identified data may be requested from the corresponding author or through the Research Ethics Office at City St George’s, University of London. Requests will be considered in accordance with participants’ consent and data protection requirements.

## Acknowledgments

We would like to thank the professionals who took part in the focus groups and interviews, the hospices that supported the study, and the research leads who supported the recruitment of participants.

## Supporting information

S1 Appendix. Study materials and practice recommendations. Sections A-G contain the topic guide, participant information, consent forms, researcher introduction, transcription notation, debrief information and practice recommendations.

S2 Checklist. COREQ reporting checklist.

## Declarations

### Authors’ Contributions

The study was designed and conceptualised by MV and RR. MV conducted the data collection and analysis. EMW provided guidance during the analysis and verified the methodological coherence of the analytic approach. MV drafted the first version of the manuscript. All authors revised the manuscript for important intellectual content and approved the final version. MV is the guarantor.

